# Whole Genome Sequencing and single-cell transcriptomics identify *KMT2D* as a potential new driver for pituitary adenomas

**DOI:** 10.1101/2024.09.17.24312241

**Authors:** Maxime Brunner, Jenny Meylan-Merlini, Maude Muriset, Sergey Oreshkov, Andrea Messina, Mahmoud Messerer, Roy Daniel, Ekkehard Hewer, Jean Phillipe Brouland, Federico Santoni

## Abstract

The pituitary gland is a main component of the endocrine system and a master controller of hormone production and secretion. Unlike neoplastic formation in other organs, Pituitary Neuroendocrine Tumors (PitNETs) are frequent in the population (16%) and, for unknown reasons, almost exclusively benign. So far, few genes have been identified as drivers for PitNETs, such as *GNAS* in somatotroph tumors and *USP8* in corticotroph tumors. Using whole genome sequencing, we uncover a potential novel driver, the histone methyltransferase *KMT2D*, in a patient in his 50s suffering from a mixed somato-lactotroph tumor. Coverage ratio between germline and tumor revealed extensive chromosomal alterations. Single-cell RNA sequencing of the tumor shows up-regulation of known tumorigenic pathways compared to a healthy reference, as well as a different immune infiltration profile compared to other PitNETs, more closely resembling the profile of carcinomas than adenomas. Genome-wide DNA methylation analysis identified 792 differentially methylated regions, including notable hypomethylation in the promoter of *SPON2*, an immune-related gene. Our results show that tumors considered as quiet and non-aggressive can share drivers, features, and epigenetic alterations with metastatic forms of cancer, raising questions about the biological mechanisms controlling their homeostasis.

## Introduction

The pituitary gland, often referred to as the master regulator of the endocrine system, lies within the sella turcica at the base of the brain. It is composed of the adenohypophysis (anterior pituitary), arising from the oral ectoderm, and the neurohypophysis (posterior pituitary), originating from the neuroectoderm. The neurohypophysis consists primarily of axonal projections from the hypothalamus. Together with the hypothalamus, the pituitary gland plays a critical role in regulating essential physiological processes such as growth, puberty, metabolism, stress responses, reproduction, and lactation through hormone production and secretion.

Pituitary tumors, or Pituitary NeuroEndocrine Tumors (PitNETs), are the third most common type of intracranial tumors, accounting for approximately 15% of cases [1]. Although many PitNETs remain undiagnosed (with an autopsy prevalence of around 16%), the incidence of clinically relevant cases in the United States was estimated at 4.07 per 100,000 per year between 2012 and 2016 [2]. The vast majority of pituitary tumors are adenomas (95%), which can sometimes be invasive, invading neighboring tissues and having a higher risk of local recurrence as well as displaying metastatic lesions but only in 0.1% to 0.2% of cases [3]. Clinically, PitNETs can present with severe symptoms due to excessive hormone secretion, inhibition of specific hormone secretion, or signs and symptoms related to an expanding sellar mass, such as visual deficits and headaches.

The most common treatment for PitNETs is transsphenoidal surgery, followed by radiotherapy and drug control for certain subtypes [4]. Additionally, immunotherapy with PD-L1 has been proposed as an alternative treatment [5]. A recent study aimed to investigate the association between PD-L1 expression and the radiological and pathological behavior of PitNETs to determine its suitability as a target in relapse cases [6].

Classification of these tumors follows the latest WHO guidelines from 2022 [7], which distinguish tumors of the anterior and posterior lobes, as well as other hypothalamic tumors. Key features used to determine the type and subtype of a tumor include transcription factors, hormones, and other biomarkers, such as low-molecular-weight cytokeratin to determine the cell of origin. For instance, somatotroph tumors are divided into densely and sparsely granulated subtypes, based on the distribution of secretory granules within the tumor. Densely granulated tumors generally secrete more hormones than their sparsely granulated counterparts. These tumors are further characterized by staining for the transcription factor PIT1 (*POU1F1*, a lineage determinant) and GH (*GH1*), along with the glycoprotein hormone *γ*-subunit (*CGA*). Additionally, cytokeratin staining shows perinuclear patterns in densely granulated tumors and more than 70% fibrous bodies in sparsely granulated subtypes.

Despite significant advancements in the accurate classification of PitNETs, their etiology remains poorly understood. Genetic studies have highlighted the role of inherited germline mutations in genes such as *AIP* in Familial Isolated Pituitary Adenoma [8], *PRKAR1A* in Carney Complex, *MEN1* in multiple endocrine neoplasia syndrome type 1 (MEN 1) [9], *CDKN1B* in MEN 4 [10], and *USP8* as an oncogene in sporadic corticotroph adenomas [11], as well as *GNAS* mutations in somatotroph adenomas [12].

In this report, we present a potential novel driver for pituitary neuroendocrine tumors: the histone-lysine N-methyltransferase 2D (*KMT2D*) gene. Germline pathogenic mutations in *KMT2D* are known to cause Kabuki syndrome, characterized by distinct facial features, growth delay, and mental retardation [13]. Somatic mutations in this gene have been implicated in various cancers, including breast cancer [14], lung squamous cell carcinoma (LUSC) [15], and B-cell-derived lymphoma [16], making it one of the most frequently altered genes in cancer. However, *KMT2D* has not previously been reported as a driver for pituitary neuroendocrine tumors.

## Results

### Patient information

In the context of a large collaborative research project, Pituitary Neuroendocrine Tumors (Pit-NETS) are fresh collected after surgery at the university hospital (CHUV) in Lausanne, Switzerland and processed for single-cell RNA sequencing (sc-RNAseq) and Whole Genome Sequencing (WGS) in our laboratory. Here, we present the case of a male patient in his 50s with a secreting mixed somatolactrotroph macroadenoma causing acromegaly and elevated IGF-1 levels. Surgical intervention successfully alleviated acromegaly symptoms and normalized IGF-1 levels. Post-operative brain MRI confirmed complete tumor resection. The pathology report on the excised tissue mentions the loss of the usual acinar architecture on reticulin staining as well as cells having a densely granulated eosinophillic cytoplasm. No apparent sign of necrosis or mitosis. Cells express PIT1 (*POU1F1*), SF1, GH and PRL (Fig. 1). Proliferation index MIB1/Ki67 is around 1%. Very weak intensity staining with PD-L1 (*CD274*) for 40% of tumoral cells. The patient is referred as patient 16.

**Figure 1:**
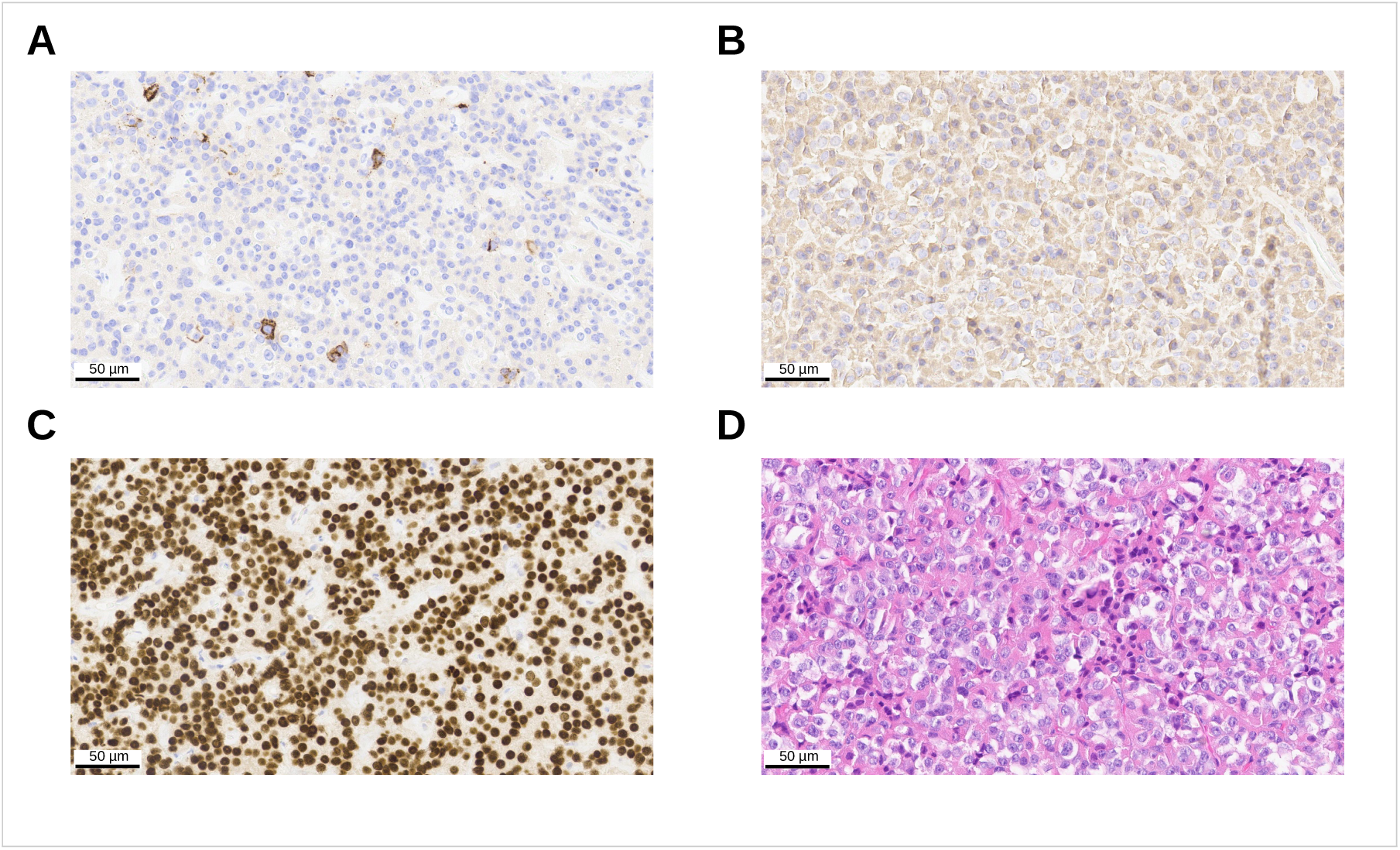
**A**: PRL staining. **B**: GH staining. **C**: POU1F1 staining. **D**: Hematoxylin and eosin staining.

### Variant information

Variant analysis of matched germline (blood) and tumor WGS from the patient revealed 455 somatic single nucleotide mutations [cf. Methods], with the only plausible candidate to be a driver in the gene *KMT2D*. The mutation is NM 003482:exon32:c.8047-2A*>*G, a splicing variant disrupting the acceptor site of exon 33. This variant is predicted as pathogenic by SpliceAI, ADA and RF [17, 18]. This variant is not reported in ClinVar and absent from gnomAD exome and genome databases. It is reported as rs2120513666 in dbSNP. Absent in germline, in the tumor tissue the variant presents with an allelic ratio of 50% indicating a tumor purity close to 100% in absence of structural variants in this region (Fig. 2A-B).

**Figure 2:**
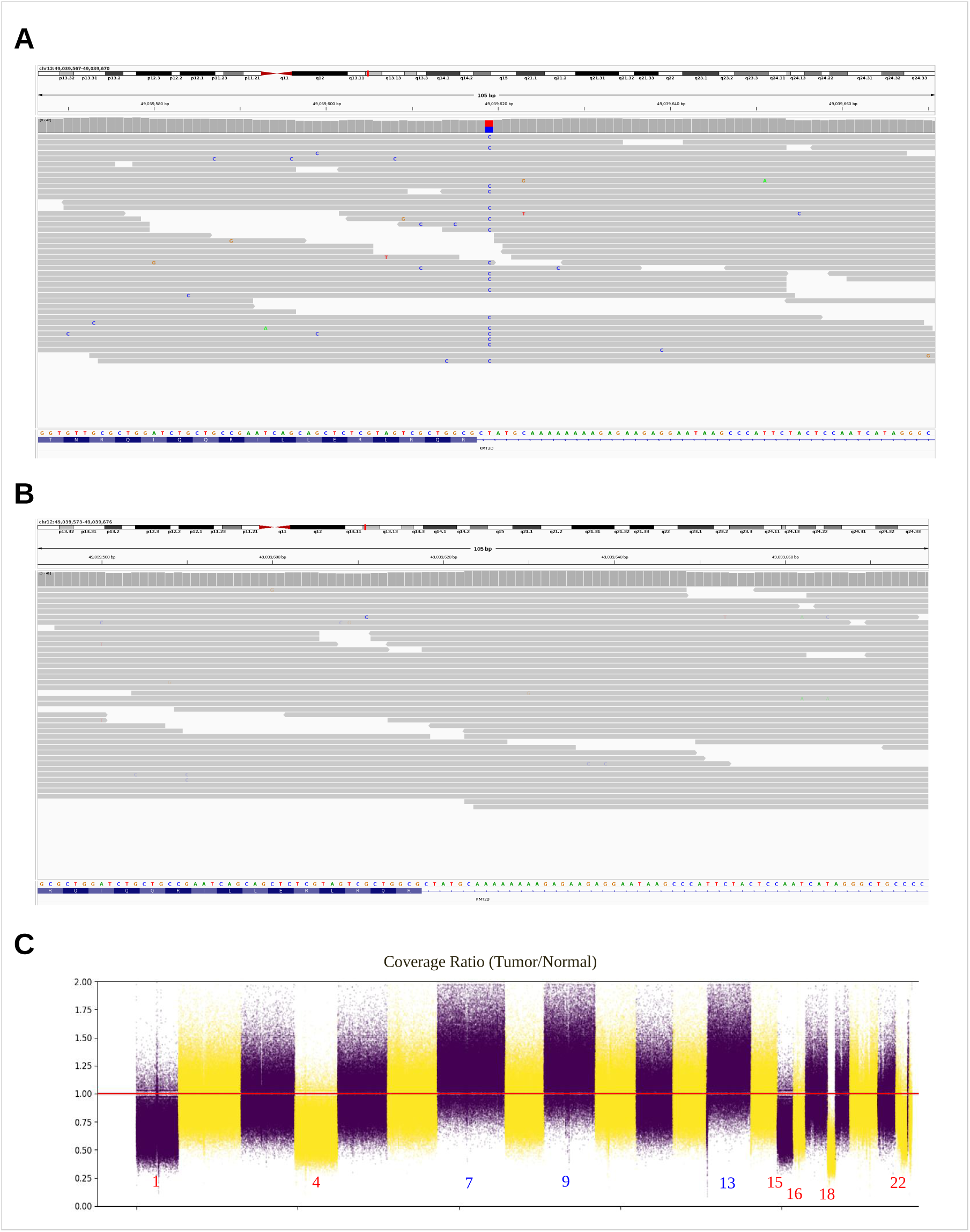
**A**: IGV output of Patient 16’s tumor WGS coverage showing the somatic mutation. **B** IGV output of Patient 16’s germline WGS coverage.**C**: log2 Matched germline and tumor coverage ratio.

### Structural alterations

Indeed differential coverage analysis [cf. Methods] between germline and tumor DNA showed extensive alteration in chromosomes 1, 4, 15, 16, 18 and 22 (full chromosomal deletions) as well as chromosomes 7, 9 and 13 (full chromosomal amplifications) but not in chromosome 12 (Fig. 2C).

### Single-cell RNA seq

Tumor gene count matrix was generated by CellRanger (version 6.1.0) and processed with Seurat for quality control, normalization, dimensionality reduction, clustering and annotation [cf. Methods]. 2928 cells were retained after stringent quality control with a mean number of detected genes of 2031 and mean unique molecular identifiers (UMIs) of 7348 per cell. Initial clustering displayed 13 clusters (Fig. 3A) which after automatic and manual annotation were separated into tumor cells (3 clusters), immune cells (6 clusters) and structural cells (4 clusters). In depth analysis enabled us to separate the tumor clusters into *GH1* and *PRL* expressing cells, the immune cells into CD4 T cells, CD8 T cells and monocytes (with subcategories, cf. Immune cell analysis) and structural cells into stem cells, pericytes, fibroblasts and endothelial cells (Fig 3B).

**Figure 3:**
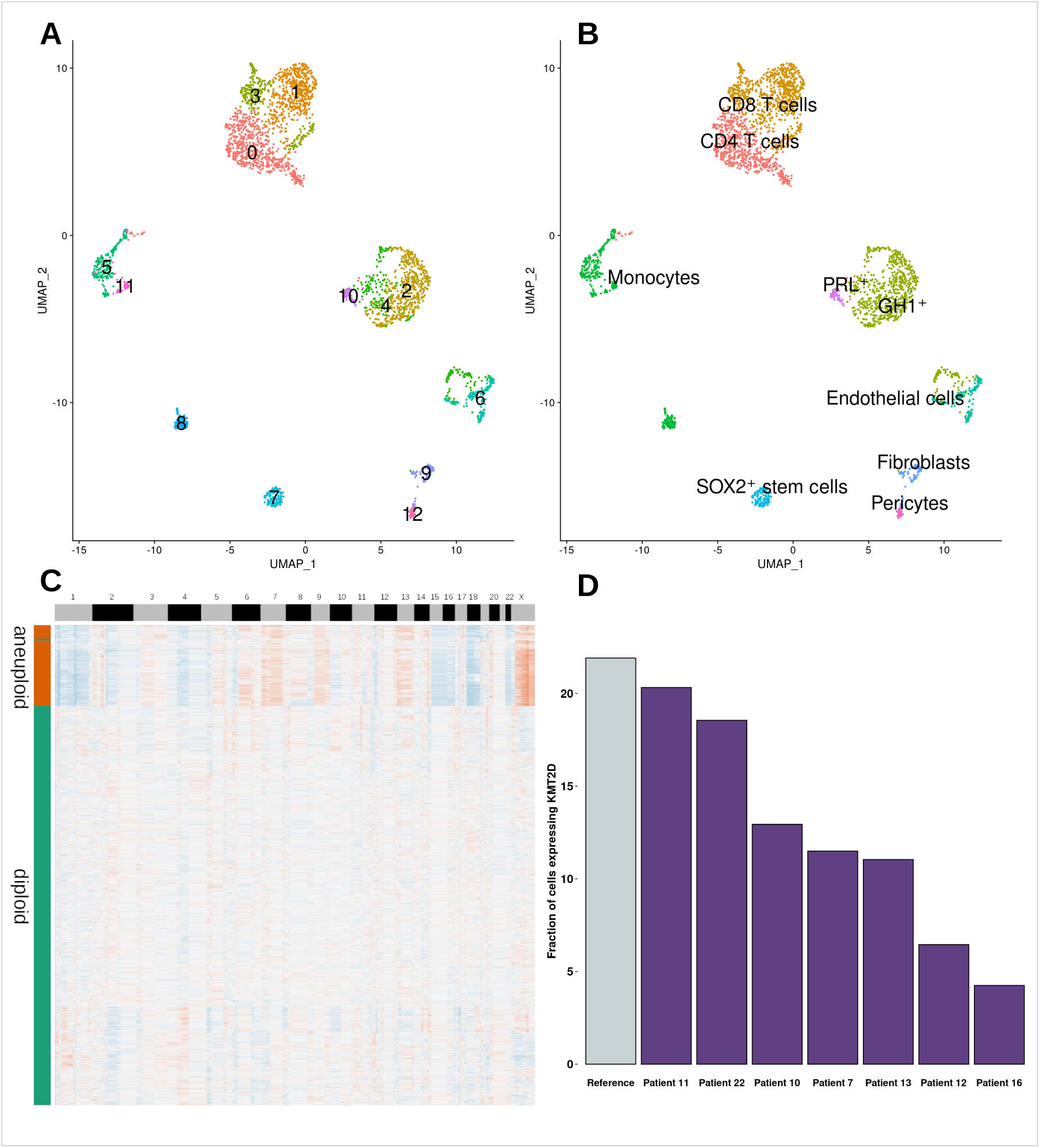
**A**: Initial clustering with 13 clusters. **B**: Annotated UMAP based on canonical markers and automated annotation pipeline. **C**: Copykat heatmap representing chromosomal alterations. Blue for deletion and orange for amplification. Y axis bar represent predicted aneuploid and diploid cells. **D**: Number of cells expressing *KMT2D* in different PitNET samples and the healthy reference.

### Alterations at single-cell level

We validated the chromosomal alterations using Copykat in single cells (version 1.1.0). This software predicts the aneuploid or diploid status of each individual cell allowing for a clear separation of tumor cells versus normal cells. The resulting heatmap showed essentially the same pattern of altered chromosomes we observed in WGS with little differences mostly due to the increased coverage in WGS with respect to single-cell transcriptomics (Fig. 3C).

### KMT2D expression in PitNETs

The fraction of cells expressing *KMT2D* was computed in all PitNET sc-RNA seq data sets and also in an integrated reference (somatotroph and lactotroph cells from the single-cell data set of 3 healthy anterior pituitary gland [19])(Fig. 3D). A consistently lower fraction was detected in patient 16 compared to all the other tumors and reference indicating a lower transcript abundance due to the non-sense mediated decay of the aberrant spliced transcript.

### Differential gene expression analysis (DGEA)

We performed a differential gene expression analysis between the integrated reference and the tumor cells using a pseudobulk approach [cf. methods]. Hallmark gene sets (H) from the Broad Institute were selected to detect up- and down-regulated pathways. Results showed up-regulation of multiple terms (Fig. 4A) which can be grouped into different categories; energy metabolism (Oxidative phosphorylation (NES: 4.18, adjusted P value: 1.26e^-23^), adipogenesis (NES: 2.39, adjusted P value: 1.1e^-4^), proliferation and cell growth (MYC targets (NES: 3.91, adj. p = 6.67e^-20^), MTORC1 signaling (NES: 2.73, adj. p = 2.16e^-20^), E2F targets (NES: 2.32, adj. p = 5.59e^-4^), DNA damage and repair mechanisms (DNA repair (NES: 2.9, adj. p = 9.61e^-7^), P53 pathway (NES: 2.4, adj. p = 3.4e^-4^), Apoptosis (NES: 2.25, adj. p = 1.51e^-3^)), stress response and survival (Unfolded protein response (NES: 2.41, adj. p = 4.33e^-4^), ROS pathway (NES: 2.33, adj. p = 7.21e^-4^)), immune response and inflammation (Interferon gamma (NES: 2.55,adj. p = 1.02e^-4^) and alpha response (NES: 2.4, adj. p = 6.19e^-4^), TNFA signaling (NES: 2.38, adj. p = 4.33e^-4^)) and cell signaling pathways (UV Response up (NES: 2.69, adj. p = 3.23e^-5^). Down-regulated pathways are KRAS signaling down (NES:-2.17, adj. p = 2.09e^-3^)) and UV Response down (NES: -2.11, adjusted P value: 2.89e^-3^)). Interestingly, the GO term ”KRAS signaling down” indicates a reduced expression of of known KRAS downregulated genes. Indeed, KRAS appears up-regulated in the tumor (log2FC: 1.47, adjusted P value: 3.34e^-35^).

**Figure 4:**
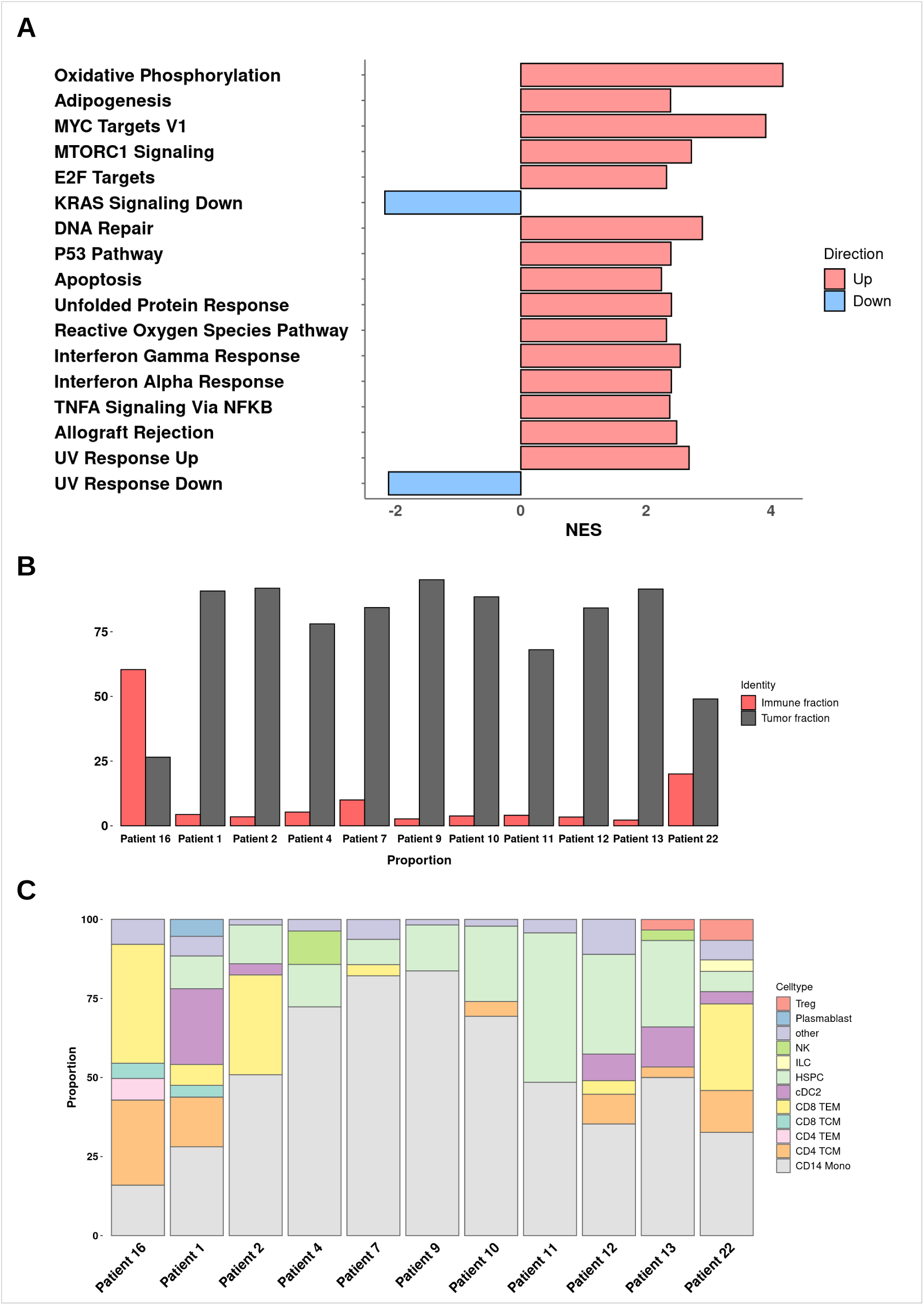
**A**: Differential gene expression analysis results showing HALLMARKS up- and downregulated between patient 16 and reference. **B**: Tumor and immune cells proportion in PitNET samples. **C**: Immune cells composition in the PitNETs predicted by Azimuth.

### Immune cells analysis

While exploring the general composition of patient 16’s sc-RNA seq, we noticed a peculiar distribution of cell-types compared to the other PitNETs (i.e. increased number of immune cells proportional to the tumor cells). In this specific single cell sample there are more than 60% immune cells while generally in PitNETs, the proportion is around 5% (Fig. 4B). To determine the composition of immune cell subcategories, we used Azimuth (https://app.azimuth.hubmapconsortium.org) for label transfer (i.e. automatic annotation) was used. It allows for the prediction of the most likely cell type compared to a comprehensive list of peripheral blood mononuclear cells (PBMCs) [20]. Predicted cell-types were effector memory CD8 T cells (37.6%), central memory CD4 T cells (26.9%), CD14 monocytes (15.9%), effector memory CD4 T cells (6.8%), central memory CD8 T cells (4.8%) and different, low proportion immune cell-types that we grouped in an ”other” category (7.9%). Then, we performed the same analysis for the other PitNETs as comparison and results showed a lower fraction of monocytes, a higher fraction of central memory CD4 T cells as well as the presence of effector memory CD4 T cells (absent in all others). In addition, there is a general increase in cytotoxic activity (both effector and central memory CD8 T cells). Lastly, there is a complete absence of hematopoietic stem and progenitor cells (HSPCs - present in all others) (Fig. 4C).

### Methylation profile in PitNETs

To investigate the epigenetic landscape of pituitary tumors, we conducted a genome-wide DNA methylation analysis across our cohort of PitNETs. We focused our analysis on the 1000 most variable methylation probes to identify significant patterns. The boxplots illustrate the methylation levels of these probes across all tumor samples in our cohort (Fig. 5). Our analysis confirmed the previously reported hypomethylation in POU1F1-PIT1 lineage [21], which includes somatotroph, lactotroph, and thyrotroph tumors. These tumors, including Patient 16’s somatotroph tumor, consistently exhibited lower overall methylation levels across the most variable probes compared to other tumor types. Notably, despite the presence of the *KMT2D* mutation and extensive chromosomal alterations in Patient 16’s tumor, we did not detect significant DNA methylation differences between this tumor and other somatotroph tumors.

**Figure 5:**
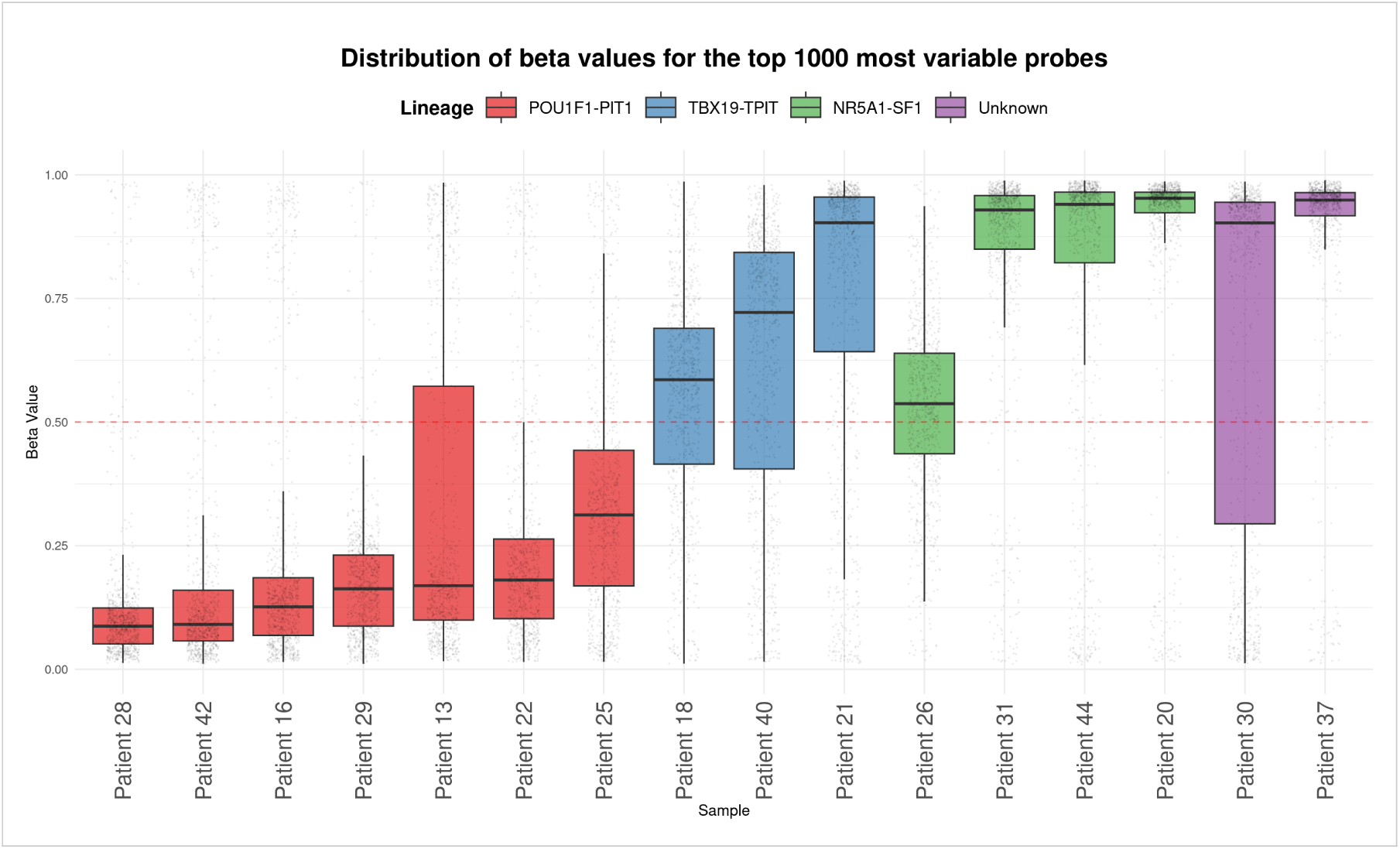
Boxplot representing beta values of the top 1000 most variable probes in 16 PitNETs. POU1F1 lineage includes somato- and lactotroph tumors. TBX19 lineage produces corticotroph tumors and NR5A1 gonadotroph tumors (Non-functioning).

## Discussion

In this case report, we present a patient in his 50s with a secreting mixed somato-lactotroph tumor of the pituitary gland exhibiting extensive chromosomal alterations. Variant calling from whole-genome sequencing (WGS) data revealed a splicing variant in *KMT2D*. This histone methyltransferase enzyme is associated with active gene transcription and chromatin regulation. In normal conditions, it acts as a tumor suppressor gene, preventing uncontrolled cell growth and division [22]. The loss of function of *KMT2D* disrupts histone methylation, leading to dysregulation of DNA damage repair and gene expression [23]. This disruption increases chromosomal instability, which can potentially lead to severe forms of cancer [24–26]. However, it has not been previously reported as a driver for pituitary neuroendocrine tumors (PitNETs). Subsequent single-cell RNA sequencing (scRNA-seq) analyses of the tumor revealed unusual results compared to other PitNETs. Firstly, a reduced number of *KMT2D* transcripts were detected compared to a healthy reference for the action of the nonsense mediated decay (NMD) pathway on the aberrantly spliced transcript. Differential gene expression (DGE) analysis showed up-regulation of several pathways associated with tumor maintenance and progression, such as oxidative phosphorylation, DNA repair mechanisms, inflammation and immune response, and KRAS signaling. This suggests similarities between this adenoma and more aggressive carcinomas . The extensive chromosomal alterations observed in this patient are the consequence of a significant chromosomal instability, which is often a hallmark of cancer progression. Chromosomal instability can lead to an increased mutation rate and the activation of oncogenic pathways. In this case, the up-regulation of KRAS related pathways is noteworthy. KRAS is a well-known oncogene, and its activation can drive tumorigenesis through various mechanisms, including promoting cell proliferation and survival, metabolic reprogramming, and evasion of immune surveillance. However, unlike in lung cancer where up-regulated KRAS disrupts the circadian gene *PER2* leading to increased activation of glycolytic pathways [27], this disruption is not observed in our patient’s tumor. This difference may be tissue-specific, indicating that the role of KRAS activation varies across different tumor types. Immune cell analysis between patient 16 and other PitNETs highlighted an inverse ratio (more immune cells than tumor cells), indicating an increased involvement of the tumor microenvironment. Moreover, immune infiltration was different, with increased immune surveillance and cytotoxic activity (higher numbers of CD4 and CD8 T cells). We speculate that this might counteract a possible transition to malignancy and that the tumor microenvironment, including immune cell infiltration, plays a significant role in preventing the adenoma from progressing to carcinoma. Immunohistochemical analysis revealed light PD-L1 expression in approximately 40% of tumor cells in this pituitary neuroendocrine tumor. While PD-L1 expression is often associated with immune evasion in various cancer types, its significance in PitNETs appears to be more nuanced. Our review of other pathology reports indicates that PD-L1 expression levels in PitNETs can be highly variable, with even some indolent adenomas showing weak expression in up to 80% of cells. Thus, PD-L1 expression alone may not be a reliable indicator of tumor aggressiveness or invasiveness in this context. However, when considered alongside other findings—such as the *KMT2D* mutation, extensive chromosomal alterations detected by WGS, and the unusually diverse immune cell infiltrate observed in single-cell analysis—it contributes to a complex picture of tumor biology. While these collective findings don’t necessarily indicate malignancy, they highlight the tumor’s unique characteristics and emphasize the need for comprehensive, multi-modal analysis in tumor characterization. Our methylation analysis further supports this complex picture of tumor biology. We analyzed the 1000 most variable methylation probes across our cohort of pituitary tumors. Interestingly, we observed that somatotroph tumors, including Patient 16’s tumor, consistently showed lower overall methylation profiles compared to the other tumor types. This hypomethylation pattern in somatotroph tumors suggests a distinct epigenetic landscape that may contribute to their unique cellular characteristics and behavior. Notably, despite the presence of the *KMT2D* mutation and extensive chromosomal alterations in Patient 16’s tumor, we did not find significant methylation differences between this tumor and other somatotroph tumors. This suggests that the *KMT2D* mutation in this context may not dramatically alter the overall methylation profile beyond what is typically observed in somatotroph tumors. Interestingly, although epigenetic alterations, along with strong drivers as *KMT2D* and significant genetic and transcriptomic changes may contribute to bridge the gap between pituitary adenomas and carcinomas, PitNETs, as in the presented case, appear resistant to malignant transitions. This suggests that the interplay between genetic mutations, epigenetic regulation, and the tumor microenvironment, in the context of the particular anatomical location outside but yet close to the brain, may be implicated in the maintenance of PitNETs as adenomas. Understanding this peculiar tumor behavior may offer new avenues for targeted therapies or diagnostic approaches in the future.

## Data availability

Single cell processed data are available in NCBI GEO under the accession number XXX.

## Author contributions

F.S designed and supervised the study, M.B., J.M. and Maude.M. performed the experiments, A.M. helped with the experiments, M.B. analysed the single cell and methylation data, S.O. and F.S. analysed the genetic data, Mahmoud.M. and R.D. provided the tumors and clinical data, H.E. and J.P.B. analysed the tissues and wrote the clinical reports, M.B. and F.S. wrote the manuscript. All authors contributed to the manuscript.

## Methods

### Tissue collection and dissociation

PitNETs are resected at the university hospital in Lausanne, Switzerland. After the operation, the department of pathology takes half of the fresh tumor to perform routine analyses and the clinical report. The other part is dedicated to this project. To dissociate the cells we are taking advantage of Worthington papain dissociation kit. In short, tumor is first minced to break the strongest adherence between tissue and cells. Then pieces are mixed with the digestion mix (EBSS, papain, DNAse and collagenase) and heat up at 37°C for 30 minutes in a shaker set to 60 RPM. Every 10 minutes, the liquid is passed through a 10 mL pipette to further separate the cells from the tissues. At the end of the incubation, remaining visible parts are mechanically dissociated with a 200µl pipette tip set on a 1000µl tip and pressed to the side of the tube to ensure maximum dissociation thank to laminar flow. Next, sample is filtered on a 40µm cell strainer and centrifuged for 5 minutes at 300g. The pellet is resuspended in 500µl diluted 1/10 ovomucoid (trypsin inhibitor) to stop the digestion process. Next steps involve gradient centrifugation in concentrated ovomucoid for 6 minutes at 70g to get rid of aggregates and dead cells. Pellet is then resuspended in 600µl of cold HBSS and filtered a second time on a 40µm cell strainer. Cells are counted before the second gradient centrifugation in order to resuspend them in the desire volume of HBSS to hit a range of 800-1200 cells per µl.

### Single-cell capture, cDNA library preparation, and sequencing

For this project, we used the 10X Chromium workflow. A Chromium Next GEM Chip G (10X Genomics) is loaded with approximately 6’000 cells and sequencing libraries are prepared strictly following the manufacturer’s recommendations (manual CG000204 revD). Briefly, an emulsion encapsulating single cells, reverse transcription reagents, and cell barcoding oligonucleotides are generated. After the reverse transcription step, the emulsion is broken, and double-stranded cDNA are generated and amplified for 12 cycles in a bulk reaction. The cDNA is fragmented, a P7 sequencing adaptor is ligated, and a 3’ gene expression library generated by PCR amplification for 12-14 cycles depending on the initial cDNA amount. Libraries were quantified using a fluorimetric method (Q-Bit), and their quality was assessed on a Fragment Analyzer (Agilent Technologies). Sequencing was performed with DNBSEQTM technology in BGI Europe (Denmark). Primary data processing and assembly was performed with the Cell Ranger Gene Expression pipeline (version 6.1.0, 10X Genomics) with GRCh38 (refdata-gex-GRCh38-2020-A) as transcriptome reference.

### DNA extraction and WGS

Extraction of genomic DNA is done with AllPrep DNA/RNA Mini Kit (Qiagen) following standard protocol. Tumor and matched blood samples have been whole genome sequenced by BGI Poland with PE150 30X/90Gb on DBNSEQ machines. Alignment and mapping has been performed by a custom pipeline based on Sentieon v202308.03 [28] with GRCh38 human genome reference. For somatic variant discovery TNScope algorithm from Sentieon [29] was used in a tumor-normal matching sample mode. From resulting variant set any variant not passing any TNScope filter was removed as potentially of non-tumor origin.. Annotation of resulting vcf files with tumor variants has been performed with Annovar annotation tool using dbSNP v150, SpliceAI, gnomad exonic v 2.11 and gnomad genomic v 3.0 databases [30].

### Match germline tumor coverage ratio

Analysis of structural variations has been performed through tumor vs matched germline (blood) coverage ratio. Germline single nucleotide variants with allelic ratio = 0.5 and quality score *>* 400 have been matched with overlapping tumor variants and the log2 ratio of the normalized coverages per variant reported.

### Sample processing

Data processing was done with R (version 4.2.1). For each samples, the same processing is used for quality control and downstream analyses. Quality control is performed using SingleCellExperiment (version 1.26.0), DropletUtils (version 1.24.0), scuttle (version 1.14.0), AnnotationDbi (version 1.66.0) and scDblFinder (version 1.18.0). Shortly, data object is created and genes are mapped to chromosomes using EnsDb.Hsapiens.v86 (version 2.99.0). Next, cells expressing less than 200 genes are removed from the analysis. Then to account for outliers in mitochondrial gene expression, isOutlier is used with default parameters (Median Absolute Deviation = 3). Afterward, to detect and remove putative doublets, scDblFinder with dbr.sd = 1 is used. Finally, the main processing was done with Seurat [31] (version 5.0.3) using high quality cells as input. Data normalization is performed with SCTransform() followed by linear dimension reduction (RunPCA(), default parameter), non-linear dimension reduction (RunUMAP(), with dims = 1:20, neighbors detection with Share Nearest Neighbor graph (SNN graph, FindNeighbors() with dims same as above) and finally cluster detection (FindClusters() with default parameters). First round of cell-type assignment was done by i. extracting markers per cluster (FindMarkers(), default parameters) and comparing them with current knowledge, ii. literature reviewing.

### Integration and differential gene expression analysis

Prior to perform differential gene expression analysis, integration of 3 data sets of healthy adult pituitary gland to create a unified data set is performed with Seurat. Briefly, individual data set previously processed with the above described workflow are merged into a single object. Then, object layers are integrated with IntegrateLayers() using RPCAIntegration as integration method. Rest of the processing is the same.

Differential gene expressions analysis is then performed in a pseudobulk because of an eccess of false positive in the default Seurat workflow [32]. Thus we used DESeq2 (version 1.44.0) package for bulk DGEA. Briefly, each data set (i.e. the integrated reference and patient 16) were split into four pseudo-replicates with an equivalent number of cells and all clusters were pulled together and considered as one single cluster. Then, DESeq2 with default parameter was applied. Genes not present in a least 20% of the cells were removed and differentialy expressed genes with adjusted p-value of *<* 0.01 and log2 fold change *>* 1 & *<* -1 are kept.

Finally, to perform gene ontology, fgsea [33] (version 1.30) was used with H: hallmark gene sets and C5: ontology gene sets from GSEA MSigDB collections from the Broad Institute (https://www.gsea-msigdb.org/gsea/msigdb/collections.jsp). Number of permutation (nPerm- Simple) was set to 10000 and pathways with an adjusted p-value *<* 0.05 are kept.

### Azimuth label transfer

For the immune cell analysis, we annotated them using Azimuth webtool and the Human PBMCs reference (https://app.azimuth.hubmapconsortium.org/app/human-pbmc). Azimuth is a set of single-cell RNA and single-cell ATAC seq references that can be used to annotate cells of a query data set. For the PBMCs reference, it contains 3 layers of increased cell-type specificity (layer 1 is more general compared to layer 2 and so on). For our analysis, we used layer 2 and plot the data using ggplot2 (version 3.5.1).

### Methylation analyis

The methylation assay was performed by the IGE3 genomics platform of Geneva University in Switzerland using Illumina Infinium Methylation Assay. In silico processing was done with the Sesame R package (1.23.8) using default parameters.

### Ethical approval

The study has been performed in accordance with the Declaration of Helsinki and approved by the Commission cantonale d’éthique de la recherche sur l’^etre humain (Vaud) ref. 2019-02033. All patients have signed the informed consent.

